# Characterization and comparison of immunity against MPXV for individuals infected with MPXV or vaccinated with modified vaccinia Ankara vaccines

**DOI:** 10.1101/2024.01.29.24301921

**Authors:** Aurélie Wiedemann, Mathieu Surénaud, Mathieu Hubert, José-Luis Lopez Zaragoza, Alexandre Ribeiro, Cécile Rodrigues, Emile Foucat, Harouna Diombera, Corinne Krief, Olivier Schwartz, Jean-Daniel Lelièvre, Yves Lévy

## Abstract

The 2022 monkeypox virus (MPXV) outbreak has revitalized questions about immunity against MPXV and vaccinia-based vaccines (VAC-V), but studies are limited. We analyzed immunity against MPXV in individuals infected with MPXV or vaccinated with the licensed modified vaccinia Ankara vaccine (MVA)-BN or an experimental MVA-HIVB vaccine. The frequency of neutralizing antibody (NAb) responders was higher among MPXV-infected individuals than MVA vaccinees. Both MVA vaccines induced similar and strong humoral responses. Similarly, we show a higher frequency and magnitude (5-fold) of T-cell responses, mainly mediated by CD8^+^ T cells, against a peptide pool containing selected sequences from MPXV, Variola, and VAC-V in MPXV-infected individuals than MVA vaccinees. We describe a hierarchy of cross-reactive T-cell responses against five peptide pools that are highly homologous between VAC-V and MPXV 2022, with the highest frequency of responders against MVA-121L and MVA-018L proteins. Both vaccines stimulated a notable frequency of polyfunctional CD4^+^ and CD8^+^ T-cell responses, with a subset of CD4^+^ T cells showing a mixed cytokine profile. Finally, we found that smallpox vaccination in childhood positively affected humoral but not T-cell vaccine responses, whereas these responses were not affected in people living with HIV. These findings contribute to deciphering and monitoring the profile of immunity to MPXV and MVA. In the context of a potential threat of the reemergence of smallpox following bioterrorism, the diversification and availability of potent vaccines is crucial. The comparable immunogenicity of both MVA vaccines emphasizes the potential utility of MVA-HIVB as a valuable new tool for controlling MPXV outbreaks.

## Introduction

Mpox virus (MPXV, formerly known as monkeypox) is a zoonotic poxvirus disease that is usually endemic to countries in West and Central Africa and caused by an Orthopoxvirus (OPXV). This viral genus includes smallpox. The cessation of massive vaccination in 1980 after the eradication of smallpox has increased the risk of the emergence of OPXV in humans. Since then, the number of cases and geographic range have increased substantially, possibly related to the waning of smallpox vaccine-induced immunity. The recent outbreak in 2022 is linked to a genotypically distinct MPXV related to West Africa. It was first detected in Europe and then spread throughout the world ^1^. The large number of cases in more than 110 countries represented a global threat. This prompted the World Health Organization to claim a public health emergency of international concern.

The recent 2022 mpox outbreak was unusual, with an absence of direct links to endemic countries and sustained human-to-human transmission. Mpox causes an illness with a febrile rash in humans, similar to but milder than smallpox. Populations at risk were people living with HIV (PLWHIV), men who have sex with men (MSM), bisexuals, and young individuals without previous smallpox vaccination during childhood ^2, 3, 4, 5^. The case fatality rate was low (0.1%) relative to previous outbreaks that occurred in West (lethality rate 1-6%) and Central Africa (11%), but infections were severe and the lethality rates were high among PLWHIV with advanced infection ^4^.

Three generations of vaccines based on vaccinia virus (VACV) were originally developed against smallpox ^6^. The first-generation vaccine is comprised of live VACV, (e.g., Dryvax) and was used for the eradication of smallpox. ACAM2000 is a second-generation vaccine, also based on live VACV (Emergent Product Development Gaithersburg, MD, USA). This vaccine offers an improved safety profile and has been made accessible to healthcare workers and individuals aged 18 years or older. During the recent outbreak, vaccination campaigns were based on a third generation of a safer vaccine consisting of an attenuated modified vaccinia virus Ankara (MVA) and is incapable of replication in humans ^7^. This vaccine, produced by Bavarian Nordic (MVA-BN, commercialized under the names of IMVANEX or JYNNEOS), is the only vaccine approved for mpox prevention. Previous studies showed these second- and third-generation vaccines to be highly immunogenic ^8, 9, 10, 11, 12^. However, data demonstrating protection by IMVANEX has only been shown in animal models ^7, 13, 14, 16^. Furthermore, this vaccine is available in only limited quantities, highlighting the need to develop other vaccine candidates to control the expansion of cases in an epidemic situation.

The recent report of new cases of MPXV reinfection in MVA-BN-vaccinated individuals ^18, 19^ is a true concern and raises questions about the protective and durable immunity conferred by this attenuated vaccine. Dryvac-induced B- and T-cell immunity can persist for years (> 50 years) ^20, 21, 22, 23, 24^. Recent studies have shown low-level and rapid waning of neutralizing antibodies after MVA-BN (IMVANEX/JYNNEOS) vaccination ^25, 26^ MVA-BN does not elicit potent maturation of the B-cell repertoire, a critical process for neutralization of the virus and durability of the response ^27^, raising the question of the respective roles of B-cell and cellular immunity in vaccine protection. Few studies have investigated T-cell responses against MPXV in humans ^28, 29, 30^. Data concerning the quality of immune responses to MPXV and the currently used third generation of MVA-based vaccines are limited ^27^.

We recently examined the neutralizing antibody (NAb) responses to MVA and MPXV in convalescent patients who had recovered from mpox infection, those who have received historic smallpox vaccination, and those who had been administered MVA-based vaccines, including an experimental MVA-HIVB vaccine ^31^. A comparison of B-cell responses showed large differences in NAb levels between the various cohorts of individuals. High NAb titers were measured in ancient smallpox-vaccinated MPXV-infected patients, suggesting potential cross-protection mediated by hybrid immunity. Here, we extended these data by characterization of the cellular immunity of individuals infected with MPXV or receiving MVA-based vaccines. We describe the cross reactivity and the impact of historical smallpox vaccination and HIV status of participants on these responses.

## Results

### Participants enrolled in the study

We analyzed peripheral blood mononuclear cells and sera from 61 individuals, including eight MPXV-infected individuals (21 days after symptoms onset and noted as being convalescent, Conv.) and 53 MVA-BN vaccine recipients. Individuals born after 1980, the data of cessation of smallpox vaccination, received two doses of the vaccine, with an interval of 28 to 35 days, whereas older individuals (born before 1980 and who received a smallpox vaccination during childhood) received a single dose according to national recommendations. The median age of MPXV-infected and vaccinated individuals was 33 [24-47] and 43 [32-56] years, respectively. All participants were male. Among them, 30/61 individuals were living with HIV (PLWHIV) (6/8 MPXV-infected and 24/53 vaccinated, all under antiretroviral treatment and with a CD4 count > 200/mm^3^). The study also included ten healthy volunteers (median age 24 [21-26], 70% male), who received two doses (separated by 8 weeks) of the MVA-HIVB, candidate MVA vaccine carrying HIV epitopes in the setting of the ANRS VRI01 clinical trial ^32^.

### Neutralizing antibodies induced by MPXV infection and MVA-BN and MVA-HIVB vaccination

We quantified the neutralizing activity of sera from MPXV-infected and MVA-vaccinated individuals. We used two assays to measure neutralization titers against MVA-GFP and an authentic clinical MPXV strain isolated from a typical lesion of a French patient infected in June 2022 grown on Vero E6 cells (GenBank accession number OQ249661), with or without complement supplementation, as recently reported by our group ^31^ (Figure 1). We detected anti-MVA NAbs in 50% of MPXV-infected individuals (median ED50: 38.4). Ninety-five percent (median ED50 of 128.8) of individuals born before 1980 and 13% (median ED50 of 15) born after were neutralizers after a single dose of MVA-BN, highlighting the importance of priming during childhood. The second dose of vaccine increased the percentage of neutralizers among younger individuals to 72% (median ED50: 201). The addition of complement increased NAb titers and the percentage of responders, with 100% neutralizers detected among MPXV-infected (median ED50: 594.2) and older vaccinated individuals (median ED50: 445.9) and 94% among the younger vaccinees (median ED50: 669.5) two weeks after full vaccination. We further assessed the immunogenicity of MVA as a vaccine platform by measuring NAb levels in sera from healthy donors, all born after 1980, who received two doses of the MVA-HIVB vaccine candidate in the VRI01 trial. Two weeks after the second dose, 90% were responders, regardless of the presence of complement (median ED50: 1465 and 460.3, with and without complement, respectively) (Figure 1A).

**Figure 1.**
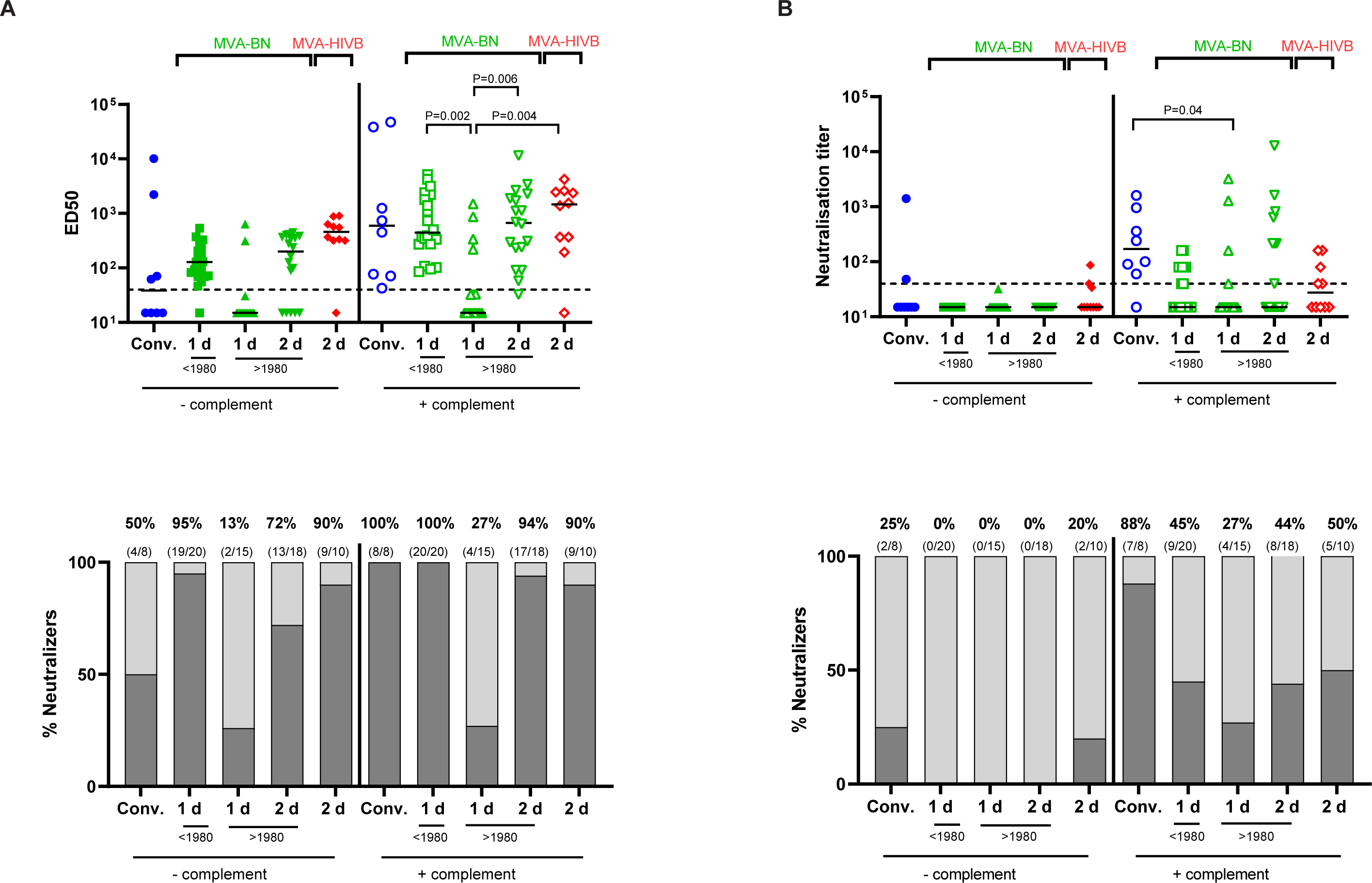
Neutralizing antibodies induced by MPXV infection and the MVA-BN and experimental MVA-HIVB vaccines. **A.** Seroneutralization of MVA-GFP and **B**. MPXV by sera from MPXV-infected patients (Conv.) (n=8) and MVA-BN- and MVA-HIVB vaccinees in the presence or absence of 10% guinea pig complement as a source of complement. Younger and older individuals were distinguished using 1980 as a cut-off year. In the vaccine groups, individuals were separated according to their vaccination status at the time of sample collection (1d: after the first dose (n=20 and n=15 for individuals born <1980 and >1980, respectively); 2d: after the second dose (n=18 and n=10, for the MVA-BN and MVA-HIVB vaccines, respectively). Anti-MVA neutralizing activity is expressed as the median effective dose (ED50), corresponding to the last dilution of plasma that reduces MVA-GFP infection by 50%. Anti-MPXV neutralizing activity is expressed as the neutralizing titer, corresponding to the highest dilution for which neutralization was observed. The dotted line represents the limit of detection (LOD). The proportion of neutralizers was evaluated by calculating the percentage of individuals exhibiting a neutralizing activity > LOD. Kruskal Wallis’ test was used for comparisons in assays performed with complement.

The NAb titers and frequency of responders were low in the absence of complement in the MPXV assay ^31^. Among MPXV-infected individuals, 25% (2/8) and 88% (7/8) were responders in assays without and with complement, respectively (Figure 1B). Among individuals who received a single MVA-BN injection (older individuals), none were responders in the absence of complement, while 45% were responders in the presence of complement. There were no responders after one or two doses of MVA-BN (younger individuals) in assays without complement, whereas 27% and 44% were neutralizers at these time points in the presence of complement. We detected 20% (2/10) and 50% neutralizers among MVA-HIVB vaccinees in assays without and with complement, respectively (Figure 1B).

### The Pan-PXV peptide pool elicits T-cell responses in MPXV-infected individuals and MVA-BN and experimental MVA-HIVB vaccinees

We next assessed T-cell immune responses using the *ex vivo* IFN-γ ELISpot assay and a commercially available pan-poxviridae peptide pool containing selected sequences from monkeypox, variola, and vaccinia viruses (Pan-PXV pool from JPT Peptide Technologies). The frequency of responders was 88% (7/8) among MPXV-infected individuals. After one dose of MVA-BN, 50% (10/20) and 53% (8/15) were responders among older and younger individuals, respectively. In the younger group, 72% (13/18) were responders after the second vaccine dose. Eighty percent (8/10) were responders after two doses of the MVA-HIVB vaccine (Figure 2A). We then compared T-cell responses specific to Pan-PXV between MPXV-infected patients and individuals who received full vaccination (1 or 2 doses, based on their history of smallpox vaccination during chidhood,) of MVA-BN, or two doses of MVA-HIVB. Infected patients showed a significantly higher median number of spot-forming cells (SFC/10^6^ cells) against the Pan-PXV pool than fully vaccinated MVA-BN individuals (540 [160-1133] vs 96.5 [22-180], P=0.0071), whereas the magnitude was not significantly different from that of recipients of MVA-HIVB (P=0.5829) (Figure 2B).

**Figure 2.**
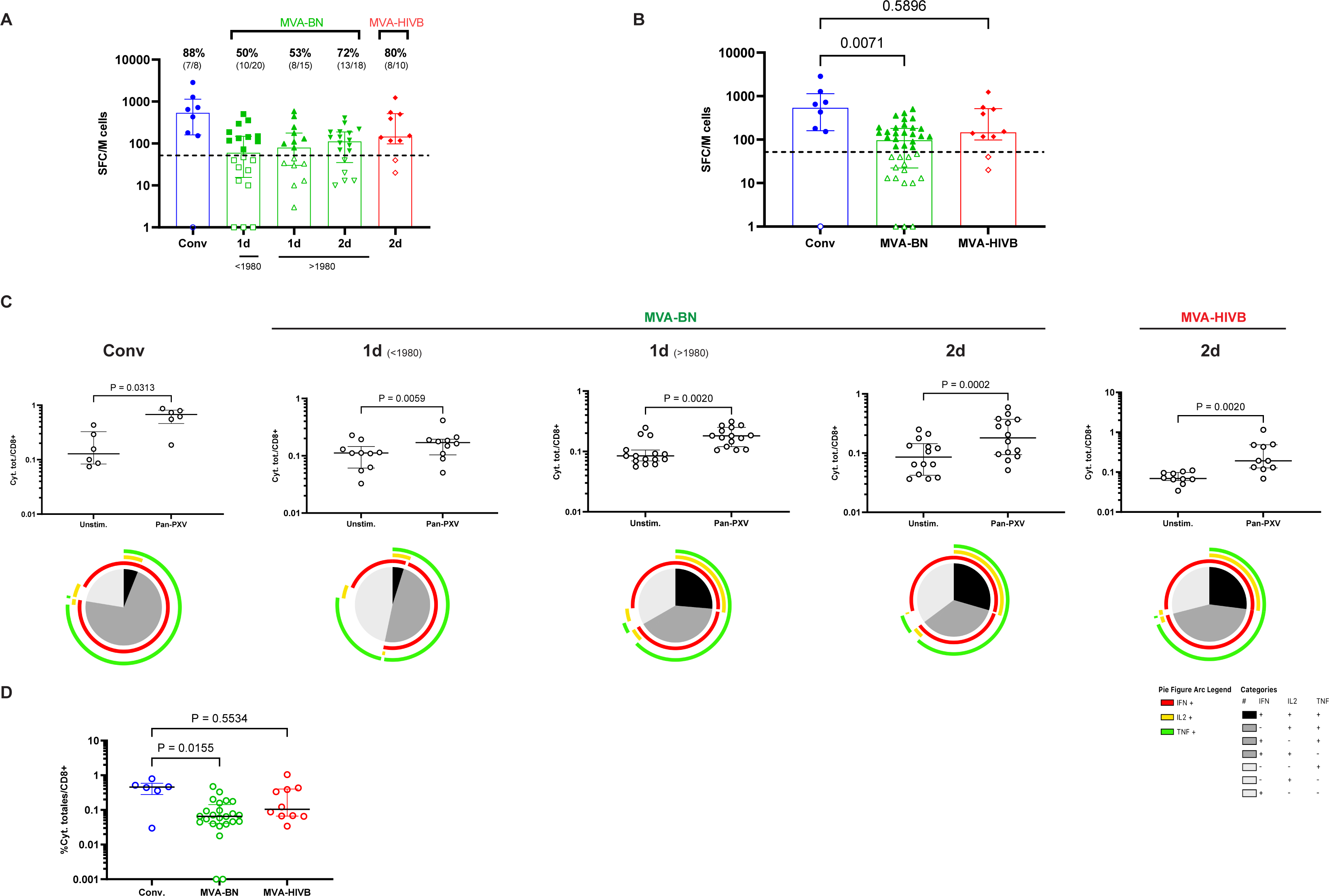
Frequency and magnitude of T-cell responses against the Pan-PXV pool of peptides induced by MPXV infection and the MVA-BN and MVA-HIVB vaccines. **A.** Magnitude of T-cell responses (SFC/10^6^ cells) of MPXV-infected patients (Conv) (n=8) and MVA-BN vaccinees (n=20 and n=15 after the first dose (1d) for individuals born <1980 and >1980, respectively, n=18 after the second dose (2d)), and MVA-HIVB vaccinees (n=10) against the Pan-PXV peptide pool measured by the IFN-γ ELISPOT assay. Background-subtracted results are presented. The dotted line represents the positivity threshold. **B.** T-cell responses (SFC/10^6^ cells) of MPXV-infected patients (Conv) (n=8), fully vaccinated MVA-BN individuals (n=38), and MVA-HIVB vaccinees (n=10) against the Pan-PXV peptide pool measured by IFN-γ ELISPOT assay. Background-subtracted results are shown, with the dotted line representing the positivity threshold. **C.** Magnitude and functional profile of Pan-specific CD8^+^ T-cell responses of MPVX-infected patients (Conv) (n=6) or MVA-BN vaccinees (n=10 and n=15 after the first dose (1d) for individuals born <1980 and >1980, respectively; n=14 after the second dose (2d)) and MVA-HIVB vaccinees (n=10) after overnight stimulation with the Pan-PXV peptide pool. The median frequency of total cytokines detected (Cyt.Tot.) (IFN-γ±IL-2±TNF)±IQR is shown. Wilcoxon comparison tests were used for statistical analysis. In the pie charts, responses are color coded according to the combinations of cytokines produced. The arcs identify cytokine-producing subsets (IFN-γ, IL-2, and TNF) within the CD8^+^ T-cell populations. **D.** Magnitude of Pan-specific CD8^+^ T-cell responses of MPXV-infected (Conv) patients (n=6), fully vaccinated MVA-BN individuals (n=24), and MVA-HIVB vaccinees (n=10) after overnight stimulation with the Pan-PXV peptide pool. The median frequency of total cytokines detected (Cyt.Tot.) (IFN-γ±IL-2±TNF) ± IQR is presented. Background-subtracted results are shown. Kruskal-Wallis and Dunn’s comparison tests were used for statistical analysis.

The functionality of Pan-PXV-specific CD4^+^ and CD8^+^ T-cell responses was assessed by intracellular cytokine staining (ICS) (Gating strategy is shown in supplementary Figure 2). We evaluated the production of IFN-γ, IL-2, TNF, and IL-4/IL-13/IL-10 (Th2/regulatory cytokines) by CD4^+^ T cells and IFN-γ, IL-2, and TNF by CD8^+^ T cells. Interestingly, only Pan-PXV-specific CD8^+^ T cells were detected *ex vivo*. The median [IQR] frequency of CD8^+^ T-cell responses was 0.67% [0.5-0.8] for MPXV-infected individuals, 0.17% [0.1-0.2] for older individuals, 0.18% [0.1-0.2] and 0.18% [0.09-0.4] for younger individuals after one or two doses, respectively, and 0.19% [0.1-0.5] for MVA-HIVB recipients (Figure 2C). These values were significantly different from those for unstimulated conditions (P<0.05) for all comparisons. Globally, the median magnitude of CD8^+^ Pan-PXV-specific T cells was higher for infected patients than for MVA-BN-vaccinated individuals (0.45% [0.3-0.6] vs 0.06% [0.04-0.14], P=0.0155) (Figure 2D). Polyfunctional CD8^+^ T-cell specific responses to the Pan-PXV peptide pool (producing up to 3 cytokines: IFN-γ± IL-2±TNF) were detectable in all groups. MPXV-infected individuals exhibited a low frequency (6%) of CD8^+^ T cells simultaneously producing three cytokines and 70% double positive IFN-γ^+^/TNF^+^ CD8^+^ T cells. Single positive CD8^+^ T cells were predominantly IFN-γ^+^. Older individuals who received one dose of MVA-BN vaccine showed a low frequency (6%) of triple positive CD8^+^ T cells, a higher frequency of double positive (IFN-γ^+^TNF^+^) (43%) CD8^+^ T cells and similar levels of single IFN-γ- and TNF-producing cells (23% for IFN-γ and 21% for TNF). Younger vaccinees exhibited clearly higher frequencies of triple- (25%) and double- (45%) positive CD8^+^ T cells after one dose of MVA-BN. After two doses of the MVA-BN or MVA-HIVB vaccines, the functional profiles of Pan-PXV CD8^+^ T cells were similar, with a high proportion of triple- (29% and 27%, respectively) and double- (35% and 44%, respectively) positive CD8^+^ T cells (Figure 2C).

### The MVA peptide pools elicit T-cell responses in MPXV-infected individuals and MVA-BN and experimental MVA-HIVB vaccinees

We next used five PepMix pools (MVA-018L, MVA-074R, MVA-093L, MVA-105L, and MVA-121L), consisting of 15-mer peptides, overlapping by 11 amino acids, covering different proteins of MVA (Supplementary Table 1), to deeply characterize T-cell immune responses against vaccinia virus and enable a comprehensive comparison between individuals vaccinated with the MVA-BN and MVA-HIVB vaccines.

The MVA proteins exhibit a high degree of similarity with both the MVA-BN and MVA-HIVB vaccines (>99%) and with the vaccinia virus Lister strain (>98%) used in historical smallpox vaccinations. These MVA proteins show significant conservation in the MPXV 2022 sequences (>94%). We first evaluated the proportion of IFN-γ responders to the MVA global peptide pool (i.e., sum of the five Pepmix peptide pools) by *ex-vivo* IFN-γ ELISPOT assay. IFN-γ responders were 75% (6/8) for MPXV-infected individuals, 60% (12/20) for older individuals who received one dose of MVA-BN, and 60% (9/15) and 83% (15/18) for younger individuals after the first and second dose of the MVA-BN vaccine, respectively. One hundred percent (10/10) in the two-dose MVA-HIVB vaccine group were responders. The median magnitude (SFC/10^6^ cells) of the response was 107.5 [39-297] for convalescent patients, 84.5 [25-195] for older individuals who received one dose, and 88 [34-236] and 221.5 [88-465] for younger individuals after one and two doses of MVA-BN vaccine, respectively. Individuals fully vaccinated with the MVA-HIVB vaccine showed the highest response (248 SFC/10^6^ cells [153-1010]) (Figure 3A).

**Figure 3.**
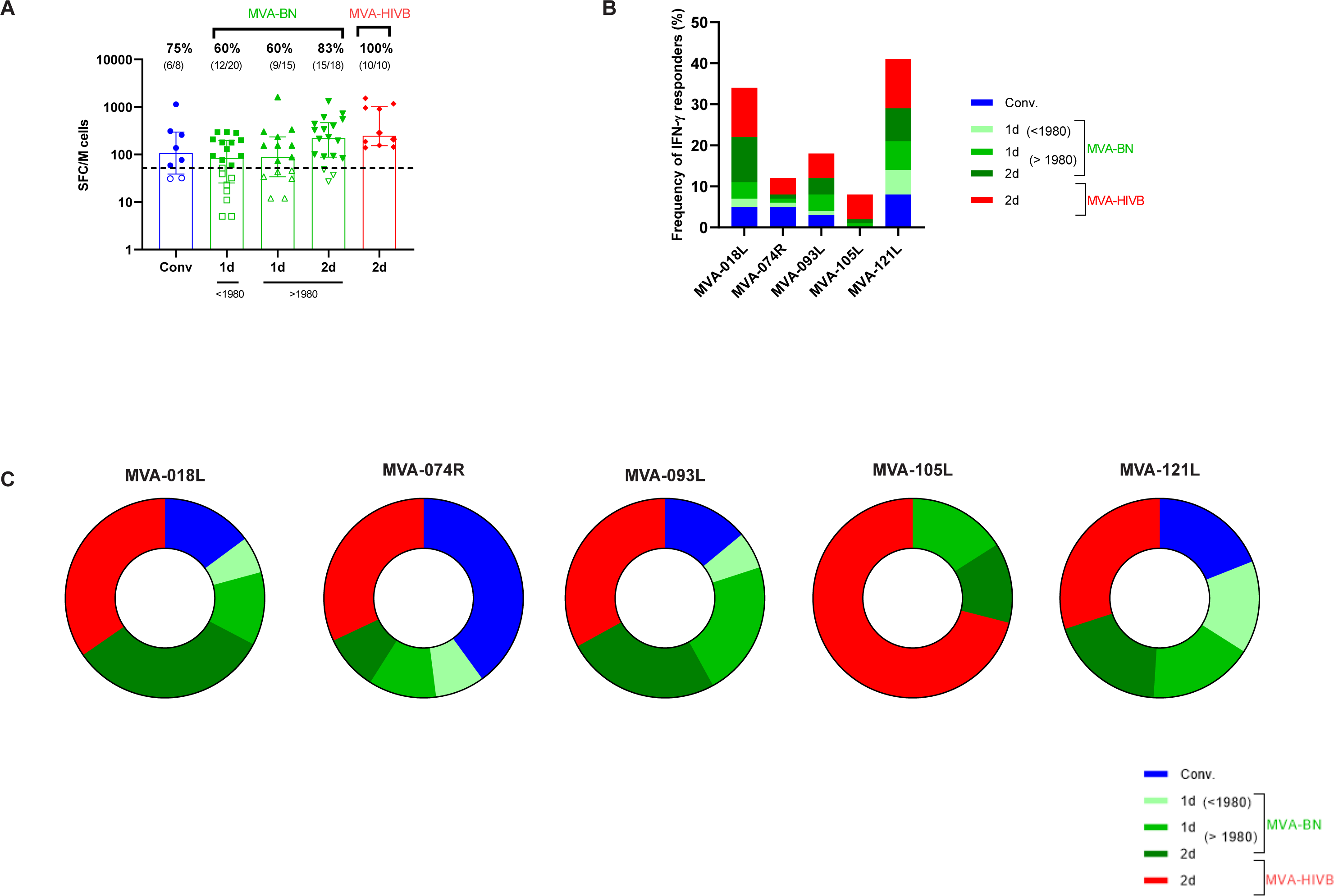
Frequency and magnitude of T-cell responses against the MVA peptide pool induced by MPXV infection and the MVA-BN and experimental MVA-HIVB vaccines. **A.** Magnitude of T-cell responses (SFC/10^6^ cells) of MPXV-infected patients (Conv) (n=8) and MVA-BN vaccinees (n=20 and n=15 after the first dose (1d) for individuals born <1980 and >1980, respectively, n=18 after the second dose (2d)) and MVA-HIVB vaccinees (n=18) against the MVA global peptide pool (sum of 5 peptide pools) measured by IFN-γ ELISPOT assay. Background-subtracted results are presented. The dotted line represents the positivity threshold. Medians ± IQR are shown. **B.** Frequency of IFN-γ responders measured by ELISPOT assay to the MVA-18L, -121L, -093L, -074R, and -105L individual peptide pools in each group of individuals. **C.** Breadth of T-cell responses against the MVA-18L, - 121L, -093L, -074R, and -105L peptide pools among IFN-γ responders.

We sought to assess the frequency of IFN-γ responders to individual peptide pools between the various groups (Figure S1). We found a higher frequency of responders to the MVA-121L and MVA-018L peptide pools (41% and 34%, respectively) than to the MVA-093L, -074R, and -105L peptide pools (18%, 12%, and 8%, respectively) (Figure 3B). We then analyzed the breadth of T-cell responses among IFN-γ responders within the five different MVA peptide pools. We found T-cell responders to each MVA peptide pool, but with differences between groups. For example, we detected T-cell responses against the MVA-105L peptide pool for younger individuals vaccinated with MVA-BN and MVA-HIVB but not for MPXV-infected and older individuals with a history of smallpox vaccination who received one dose of MVA-BN. Moreover, the MVA-105L peptide pool was mostly targeted by individuals who received the MVA-HIVB vaccine (71%). The MVA-018L and MVA-093L peptide pools were mainly targeted by individuals who received two doses of either the MVA-BN (33% and 25% respectively) or MVA-HIVB (35% and 33%, respectively) vaccines. The MVA-074R peptide pool was mostly targeted by MPXV-infected individuals (40%). Finally, responses against the MVA-121L peptide pool were equally detected for all groups (Figure 3C).

### The MVA-121L and MVA-018L peptide pools elicit polyfunctional CD4^+^ and CD8^+^ T-cell responses in MVA-vaccinated recipients

We further characterized the CD4^+^ and CD8^+^ T-cell responses induced by the MVA peptide pools by ICS (Figure 4 and supplementary figure 2 for representative dot plots). Responses were detected solely for vaccinees (MVA-BN and MVA-HIVB) and were dominated by two peptide pools: MVA-121L and MVA-018L. CD4^+^ T-cell responses to the MVA-121L peptide pool were significant (P<0.05 for all comparisons with non-stimulated conditions) for older (one dose) and younger (one and two doses) MVA-BN and MVA-HIVB (two doses) vaccinated individuals, with a high proportion of polyfunctional CD4^+^ T cells with a mixed cytokine profile simultaneously producing up to four cytokines (IFN-γ±IL-2±IL-4/IL-13/IL-10±TNF). Polyfunctional CD4^+^ T-cell frequencies ranged between 60% for younger individuals who received one dose of MVA-BN to 76% for the MVA-HIVB vaccinees. Interestingly, the functional profile of CD4^+^ T-cell responses against MVA-121L was similar for recipients of two doses of MVA-BN or MVA-HIVB. Among specific CD4^+^ T cells producing only one cytokine, CD4^+^TNF^+^ cells were predominant (Figure 4A). Two doses of MVA-BN and MVA-HIVB vaccines elicited MVA-121L-specific CD8^+^T cells (P=0.02 and P=0.004 compared to unstimulated conditions, respectively), with mostly IFN-γ^+^ single positive CD8^+^ T cells (Figure 4B). In addition to MVA-121L, MVA-018L also induced detectable CD8^+^ T-cell responses following two doses of the MVA-BN and MVA-HIVB vaccines (P=0.0009 and P<0.0001 compared to unstimulated conditions, respectively), with a predominant proportion of polyfunctional CD8^+^ T cells simultaneously producing up to three cytokines (47% and 66% for MVA-BN and MVA-HIVB, respectively). Among specific CD8^+^ T cells producing two cytokines, the profile was similar for both groups, consisting mostly of IFN-γ/TNF-producing cells. The frequencies of IFN-γ^+^ single-positive CD8^+^ T cells were 49% for the MVA-BN group and 28% for the MVA-HIVB group (Figure 4B).

**Figure 4.**
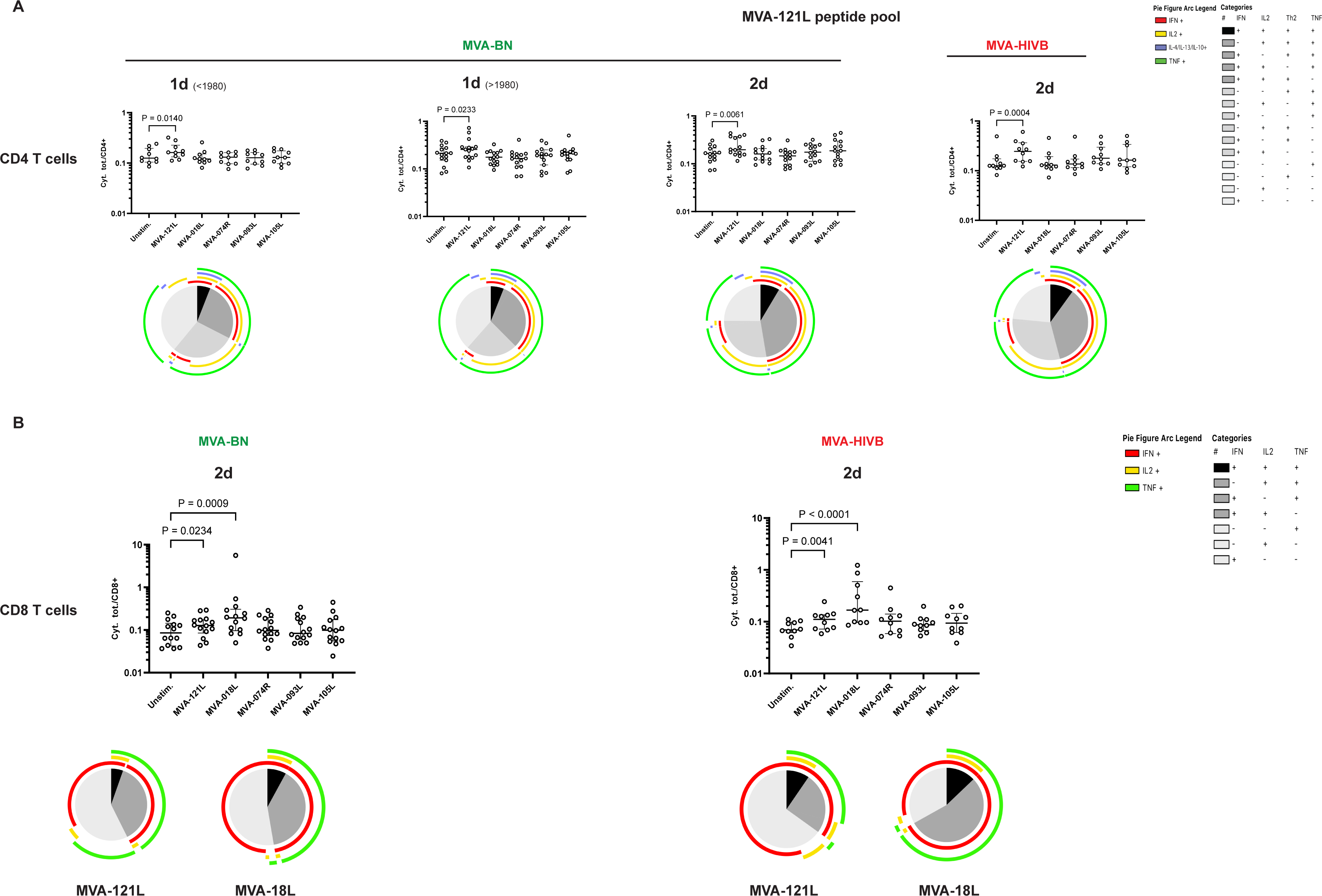
Polyfunctional CD4 and CD8 T-cell responses against the MVA-121L and MVA-018L peptide pools induced by MVA vaccination. **A.** Magnitude and functional composition of MVA-121L-specific CD4^+^ T-cell responses of MVA-BN and MVA-HIVB vaccinees. **B.** Magnitude and functional composition of MVA-121L- and MVA-018L-specific CD8^+^ T-cell responses of MVA-BN and MVA-HIVB vaccinees (n=10 and n=15 after the first dose (1d) for individuals born <1980 and >1980, respectively and n=14 after the second dose (2d)) for MVA-BN vaccinees; and n=10 for MVA-HIVB vaccinees) after overnight stimulation with five pools of overlapping peptides covering different regions of MVA. The median frequency of total cytokines (Cyt.Tot.) (IFN-g±IL-2±TNF) ± IQR is shown. Friedman and Dunn’s multiple comparison tests were used for statistical analysis. In the pie charts, responses are color coded according to the combinations of cytokines produced. The arcs identify cytokine-producing subsets within the CD4^+^ T-cell populations (IFN-γ, IL-2, IL-4/IL-13/IL-10, and TNF) and CD8^+^ T-cell populations (IFN-γ, IL-2, and TNF).

### MVA-BN vaccination in HIV-positive and HIV-negative Individuals

Finally, we evaluated the immune response of PLWHIV under successful antiretroviral therapy to MVA-BN vaccination. We found NAb titers against MVA and MPXV, as well as T-cell responses against the Pan-PXV and MVA peptide pools, for fully MVA-BN-vaccinated PLWHIV to not be significantly different from those of non-HIV-infected individuals, either after one (for older individuals) or two doses (for younger individuals) (Figure 5).

**Figure 5.**
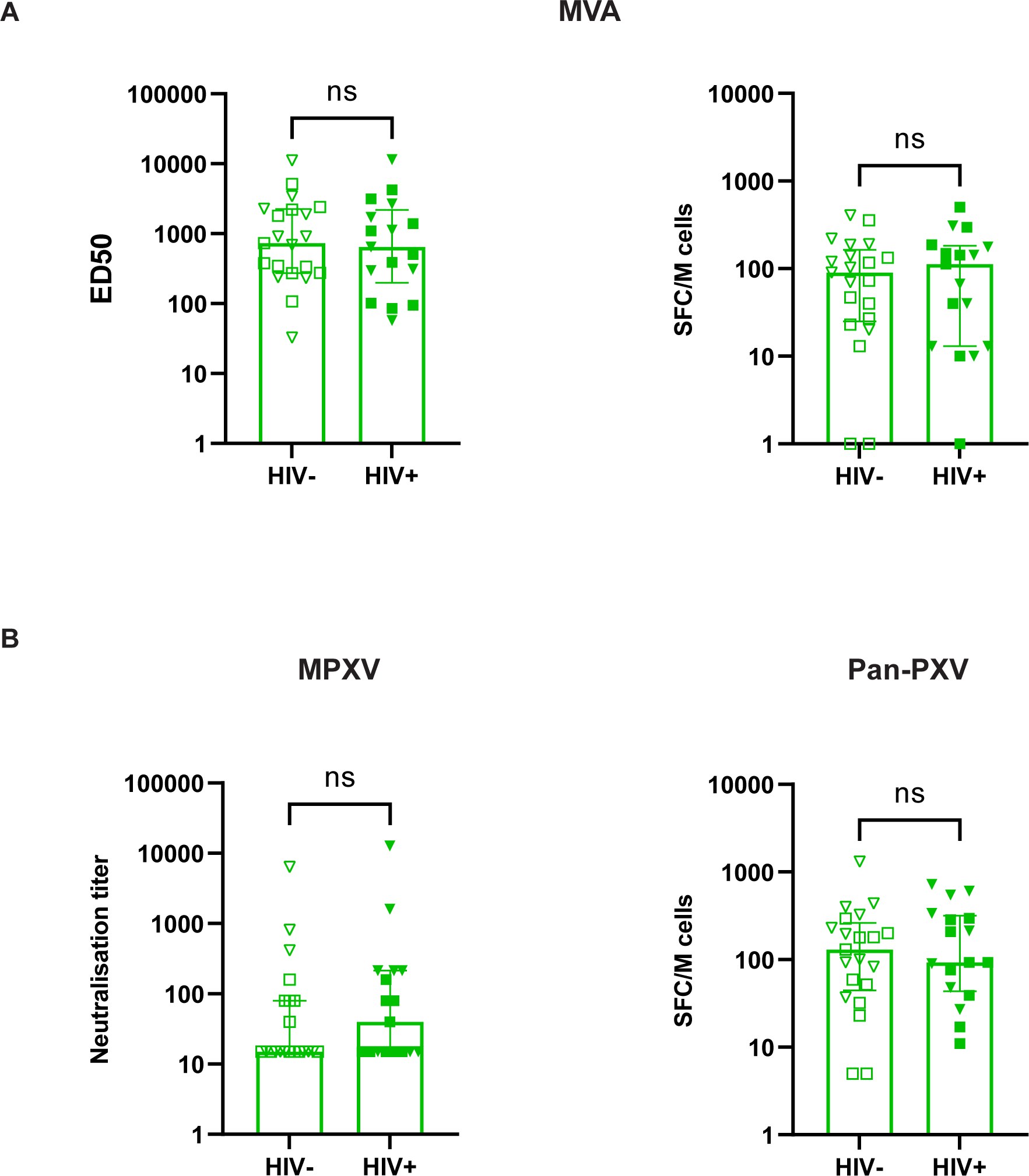
Neutralizing antibodies and T-cell immune responses elicited by MVA-BN vaccination in HIV-positive and HIV-negative Individuals. **A.** Seroneutralization of MVA-GFP (left panel) by sera from HIV^-^ and HIV^+^ MVA-BN vaccinees in the presence of 10% guinea pig complement as a source of complement. The anti-MVA neutralizing activity is expressed as the median effective dose (ED50), corresponding to the last dilution of plasma that reduces MVA-GFP infection by 50%. Magnitude of T-cell responses (SFC/10^6^ cells) (right panel) of HIV^-^ and HIV^+^ MVA-BN vaccinees against the MVA peptide pool measured by IFN-γ ELISPOT assay. Background-subtracted results are presented. **B** Seroneutralization of MPXV (left panel) by sera from HIV^-^ and HIV^+^ MVA-BN-vaccinees in the presence of 10% guinea pig complement as a source of complement. Anti-MPXV neutralizing activity is expressed as the neutralizing titer, corresponding to the highest dilution for which neutralization was observed. Magnitude of T-cell responses (SFC/10^6^ cells) (right panel) of HIV^-^ and HIV^+^ MVA-BN vaccinees against the Pan-PXV peptide pool measured by IFN-γ ELISPOT assay. (HIV^-^ n=21, HIV^+^ n=17, squares: individuals born before 1980 receiving one dose of MVA-BN, triangles: individuals born after 1980 receiving two doses of MVA-BN). Medians ± IQR are shown. The Mann-Whitney test was used for comparisons.

## Discussion

The recent outbreak of mpox infection has revitalized questions about the quality and durability of humoral and cellular responses against OPXV and the efficacy of the approved MVA-based vaccine (IMVANEX) deployed since May 2022 for populations at risk of infection. We analyzed antibody and cellular responses against MPXV and MVA in subjects infected during the recent mpox-2022 outbreak or vaccinated with the MVA-BN (IMVANEX) vaccine or an experimental MVA-HIVB vaccine (ANRS/VRI01 trial; NCT02038842) ^32^. We provide insights about the cross reactivity of the humoral and cellular responses induced by MVA-based vaccination and MPXV infection and the impact of historical smallpox vaccination on cellular immunity following MPXV infection or MVA vaccination, in particular among PLWHIV.

To study protective antibody-based immunity, we measured Nabs against MVA and MPXV. As recently shown ^31, 33^, the presence of complement enhanced the sensitivity of Nab detection. We found a higher frequency of MPXV and MVA responders among MPXV-infected individuals (88-100%) three weeks after the onset of symptoms than among individuals vaccinated with MVA. The proportion of MVA neutralizers among the recipients of two doses of MVA-HIVB or MVA-BN was roughly similar (90% and 94%, respectively), whereas the MVA Nab titers were approximately 2.2-fold higher after the second dose of MVA-HIVB than that of MVA-BN. Despite homology between MVA and MPXV, only around 45% of fully vaccinated older and younger MVA-BN-vaccinated individuals exhibited weak MPXV neutralization activity, a frequency similar to that for MVA-HIVB vaccinees (50%). Thus, despite potential differences in the MVA-BN and MVA-HIVB strains, preparation methods, and doses administered, these two vaccines elicited strong and similar humoral responses against MVA and MPXV. This observation supports the development of future MVA-based vaccines against MPXV ^34^.

One dose of MVA-BN led to a higher frequency of MVA neutralizers and higher titers (8.5-fold) for individuals born before 1980 than those born after (95% versus 26%), underscoring the importance of childhood smallpox vaccination. Smallpox vaccination of the French population stopped in 1979 and smallpox was declared to be eradicated worldwide in 1980. Our observation suggests that MVA-BN recalled long-term memory B-cell responses and supports the persistence of smallpox-specific B cells in the absence of re-exposure to smallpox antigens. Several studies have demonstrated the presence of long-lived smallpox-specific B cells up to 50 to 75 years after vaccination ^22, 35, 36, 37^ and the stability of anti-vaccinia antibody serum titers in humans for more than 80 years after first vaccination ^17, 20, 37^ Finally, the significant increase in the rate of MVA (from 26% to 72%) and MPXV (0% to 44%) responders following a second boost of MVA-BN among younger individuals reinforces the need of a complete vaccination scheme to improve protective immunity in a “naïve” population.

In humans, it is likely that both humoral and cell-mediated immunity are important for protection against OPXV. Few studies have investigated or compared cellular immune responses to MPXV- and MVA-based vaccines ^27, 28^ and more in-depth analyses of the antigens recognized by T cells are warranted. In parallel to neutralizing antibody responses, we found a higher frequency and magnitude (5-fold) of T-cell responses against peptide pools containing selected sequences from MPXV, variola, and vaccinia viruses (Pan-PXV) for MPXV-infected than MVA-vaccinated individuals. ICS assay showed that this response was mediated by CD8 T cells. This observation differs from those of a recent study ^27^ and can be explained by two principal factors. First, the cellular assays used; Cohn and colleagues adopted a combined AIM/ICS assay while we employed a two-step approach, starting with an IFN-g ELISpot and subsequently using ICS assays, despite the latter being less sensitive in comparison to an AIM assay. Our aim in employing this strategy was to deeply characterize the functionality of mpox-specific CD4 and CD8 T cells, rather than solely focusing on assessing the frequency of these cell populations. Secondly, the composition of the peptide pools, they used 2 megapools (one megapool of 238 CD8 T-cell epitopes, of which 82% are 100% conserved in MPXV, and another megapool of 300 CD4 T-cell epitopes, of which 94% are 100% conserved in MPXV). In contrast, we employed a Pan-PXV peptide pool containing 127 epitopes, comprising 107 CD8 T-cell epitopes and 20 CD4 T-cell epitopes, with 96% exhibiting complete conservation in MPXV and 94% in MVA.

In contrast to antibody responses, the frequency of T-cell responders following the administration of MVA-BN was found to be the same for both older and younger vaccinated individuals (approximately 50%). This suggests that the impact of smallpox vaccination in childhood on T-cell responses may be relatively low. Analysis of the quality of Pan-PXV specific T cells showed these responses to be mainly mediated by CD8^+^ T cells and confirmed a higher frequency of such T-cell responses among MPXV-infected individuals. Interestingly, MPXV infection was characterized by a high frequency of CD8^+^ cells simultaneously producing TNF/IFN-γ, whereas full vaccination with IMVANEX and MVA-HIVB induced a higher frequency of polyfunctional Th1 CD8^+^ T cells simultaneously producing three cytokines (IL- 2+IFN-γ+TNF).

Despite high genetic similarity between MVA and MPXV-2022 ^28, 38^, the potential impact of variations in T-cell epitopes on the cross reactivity induced by MVA-based vaccination and MPXV infection is yet to be determined. To analyze the cross-reactivity of T-cell responses in more depth, we selected five pools of peptides spanning five highly homologous regions between the two MVA vaccines and the original vaccinia Lister strain used in historical smallpox vaccination (>98%) and MPXV 2022 sequences (>94%). Using the Elispot-iFN-γ assay as a screening test, we found a hierarchy of T-cell responses against these peptide pools ranging from MVA-121L and MVA-018L (41% and 34% responders, respectively) to MVA-105L (8% responders). Indeed, the MVA-121L protein corresponds to the A10L vaccinia protein and is known to contain a high number of T-cell epitopes (27 CD4^+^ and 1 CD8^+^ T-cell epitopes) ^28, 38^, which are highly conserved (100%) in MVA-BN (21 epitopes) and MPX-2022 (25 epitopes). MVA-018L corresponds to the C7L vaccinia protein and contains two CD8^+^ T-cell epitopes ^28, 38^, including one (KVDDTFYYV) that is relatively dominant in HLA-A2^+^ individuals ^39^ The observed lower frequency of responders to MVA-18L among MPXV-infected patients relative to MVA-vaccinated individuals can be explained by the presence of the D77Y mutation in this dominant epitope in the MPXV-2022 sequence. Similarly, the frequency of T-cell responders to the MVA-093L peptide pool, which targets the H3L vaccinia protein, among MPXV-infected individuals was low, despite the presence of eight CD4^+^ and four CD8^+^ T-cell epitopes ^28, 38^. However, nine of these 12 epitopes contain at least one mutation in the MPXV-2022 sequence. Finally, T-cell responses to the MVA-105L (corresponding to the D8L vaccinia protein) peptide pool were infrequent among MVA-vaccinated individuals (8%) and not detected among MPXV-infected subjects. This is consistent with a single described CD4^+^ T-cell epitope in this protein ^28, 38^, which contains two mutations in the MPXV-2022 sequence. Overall, although the peptide regions exhibit significant homology, the variations observed among different groups of individuals align with the distinctions observed between the evolution of MVA and MPXV. Beyond a descriptive profile of T-cell responses, these data help to enrich the arsenal of peptide tools for the future immunological evaluation of OPXV infection and vaccination.

The potential threat of the reemergence of smallpox following an act of bioterrorism or of MPXV raises the question of the diversification and availability of potent vaccines. We took the opportunity to analyze B- and T-cell responses against the MVA vector of participants who received two doses of an MVA-based HIV vaccine candidate (MVA-HIVB) in the ANRS VRI01 trial ^32^. We show ^31^ and confirm that the MVA-HIVB vaccine elicits somewhat higher levels of anti-MVA Nabs in individuals born before 1980 than after IMVANEX administration. Moreover, we extend these data, showing that both vaccines induce a high frequency of polyfunctional CD4^+^ and CD8^+^ T-cell responses against MVA-121L and MVA-018L, with a high proportion of polyfunctional CD4^+^ T cells with a mixed cytokine profile simultaneously producing up to four Th1/Th2 cytokines, a population of cells described in smallpox vaccination ^40^. Overall, the T-cell responses triggered by two doses of MVA-HIVB were either similar or occurred at a higher frequency than those to MVA-BN. These findings provide reassurance concerning the potential production of clinical batches of this vector, if needed.

Although our study was not designed to compare the potency of vaccination between different groups of patients, we were able to show that Nab titers against both MVA and MPXV, as well as T-cell responses against the Pan-PXV and MVA peptide pools, were not impaired in PLWHIV, regardless of age. It’s worth noting that individuals infected with HIV-1 who participated in this study were undergoing antiretroviral therapy and maintained a robust immune profile, characterized by CD4 T lymphocyte counts exceeding 200 cells/mm3 and an undetectable plasma HIV RNA viral load (below 20 copies/ml). This favorable immunological status may account for the absence of differences in vaccine response magnitude when compared to individuals without HIV infection. Although reassuring, these data should be viewed in light of the recent observation of MPXV reinfection in previously vaccinated individuals and underscore the need to repeat boosting according to recent recommendations.

This study had several limitations. Individuals were enrolled after infection (three weeks, on average, after symptoms) and following vaccination. We did not evaluate their baseline immune status. However, the results within each group were homogeneous and allowed statistical comparisons between groups. We evaluated the levels of neutralizing antibodies, but we did not assess the levels of antigen-binding antibodies. Additionally, we did not evaluate antibody effector functions due to the unavailability of standardized kits for conducting these specific assays. We describe T-cell responses against peptides pools but did not perform epitope mapping because of the limited amount of the blood samples and lack of patient HLA information. We assessed T-cell responses using ex vivo stimulations. The lack of response in some cases may be attributed to the lower sensitivity of the ex vivo ICS assay. However, we also conducted an ELISPOT assay, which has a high level of sensitivity, even though it cannot distinguish between CD4 and CD8 responses. Finally, we did not perform long-term follow up of B- or T-cell responses to assess the durability of the responses, either for MPXV-infected individuals or MVA-vaccinated individuals.

## Data availability

All data supporting the findings of this study are available within the paper and its Supplementary Information. Raw data are available from the corresponding author upon reasonable request

## Acknowledgments

The authors thank the individuals and patients who participated in this study and Véronique Rieux (ANRS-MIE) and Lucie Hardel for their help with the ANRS-VR01 trial samples.

## Contributors

Yves Lévy, Aurélie Wiedemann, and Mathieu Surénaud conceived and designed the study. Yves Lévy, Aurélie Wiedemann, Mathieu Surénaud, Mathieu Hubert, Jean-Daniel Lelièvre, and Olivier Schwartz analyzed and interpreted the data. Mathieu Hubert, Alexandre Ribeiro, Cécile Rodrigues, Emile Foucat, and Corinne Krief performed the experiments. José-Luis Zaragoza and Harouna Diombera participated in sample and clinical data collection. Yves Lévy, Aurélie Wiedemann, and Mathieu Surénaud drafted the first version and wrote the final version of the manuscript. All authors approved the final version.

## Materials and Methods

### Participants

In July 2022, a prospective, monocentric cohort clinical study enrolling MPXV infected patients and individuals vaccinated with a third-generation MVA-based vaccine (IMVANEX) was initiated by the Immunology Department of the Henri Mondor Hospital. The study protocol was approved by the appropriate ethics committee (CPP Ile-de-France VII and IX), with approval reference 10–023 (infected patients) and ID RCB 2018_A01610-55 – n°CPP:18.09.09 (vaccinated patients). Written informed consent was collected from each participant at enrollment. In addition, we used samples from subjects enrolled in the ANRS VRI01 clinical trial, a trial performed in 2014 on 92 individuals to evaluate the effectiveness of MVA-HIVB as a vaccine candidate. Each participant was injected with two doses of MVA-HIVB with an eight-week interval. The protocol was approved by an ethics committee (Comité de Protection des Personnes Ile-de-France V, Paris, France) and the competent French health authority (Agence Nationale de Sécurité du Médicament et des Produits de Santé) and was conducted in accordance with the declaration of Helsinki. All volunteers provided written and signed informed consent for the trial. The trial was registered with ClinicalTrials.gov (NCT02038842) and EudraCT (2012-002456-17) ^32^.

### MVA and MPXV neutralization assays

Seroneutralization assays against MVA and MPXV were performed as previously described ^31^. Briefly, a modified Vaccinia virus Ankara carrying a GFP reporter gene (MVA-GFP) was provided by ANRS-MIE. A MPXV strain (MPXV/2022/FR/CMIP) was isolated from a pustular lesion of a 36-year-old French man who consulted at the Medical Center of the Institut Pasteur (CMIP), in June 2022. For the MVA neutralization assay, Vero E6 cells were used. The indicated concentrations of MVA-GFP were mixed (ratio 1:1) with serial dilutions (from 1/30 to 1/30,000) of previously heat-inactivated (30 min at 56°C) plasma or serum in the presence or absence of 10% guinea pig serum as a source of complement (GPC; Rockland). After incubation for 2 h at 37°C, the mixture was added to Vero E6 cell monolayers. Twenty hours later, cells were stained with Hoechst (1:1,000 dilution; Invitrogen). Images were acquired using an Opera Phenix high-content confocal microscope (Perkin Elmer). The GFP area and number of nuclei were quantified using Harmony software (Perkin Elmer). The percentage of neutralization was calculated using the GFP area as the value with the following formula: 100_×_(1_−_(value for serum_−_value for “non-infected”)/(value for “no serum”_−_value for “non-infected”)). The neutralizing activity of each plasma or serum sample was expressed as the ED50 (effective dose inhibiting 50% of infection). ED50 values were calculated using a reconstructed curve with the percentage of neutralization at the different serum concentrations. For the MPXV neutralization assay, U2OS cells were used. The indicated concentrations of MPXV were mixed (ratio 1:1) in a BSL-3 facility with serial dilutions of previously heat-inactivated (30 min at 56°C) plasma or serum in the presence or absence of 10% GPC. Forty-eight hours later, cells were immunostained for MPXV antigens with rabbit polyclonal anti-VACV antibodies (PA1-7258, Invitrogen) and an Alexa Fluor 488-coupled goat anti-rabbit antibody (Invitrogen). Nuclei were stained with Hoechst. Images were acquired using an Opera Phenix high-content confocal microscope (PerkinElmer). The MPXV^+^ area and number of nuclei were quantified using Harmony software (PerkinElmer). The neutralization titer was determined as the highest plasma or serum dilution in which the MPXV^+^ area was inferior to that of the “no serum” condition. The proportion of neutralizers was then evaluated by calculating the percentage of individuals exhibiting a neutralizing activity > LOD (limit of detection).

### Specific antigens

To evaluate the anti-Vaccinia and anti-Monkeypox virus cellular immune responses, screening experiments (IFN-γ ELISpot) were performed using six peptide pools from Vaccinia virus strain Ankara (PepMix MVA-018L, MVA-074R, MVA-093L, MVA-105L, MVA-121L, and MVA-189R) and one from Vaccinia virus strain Western Reserve (PepMix VACV-L3L). These pools consist of 15-mer peptides, overlapping by 11 amino acids, covering different proteins of the Vaccinia virus. In addition, the Pan-Poxviridae Select PepMix, containing 127 selected epitopes (composed of 107 CD8 T-cell epitopes and 20 CD4 T-cell epitopes, of which 96% are 100% conserved in MPXV and 94% are 100% conserved in MVA) from the Monkeypox virus, Variola virus, and Vaccinia virus, was used. The peptide pools were all purchased from JPT Peptide Technologies, Berlin, Germany. As the sequence homology of Putative 21.7k protein (MVA189R) was below 90% between MVA and MPXV and as no responder was observed with VACV-L3L in screening experiments, these two peptide pools were not used in later experiments. The five remaining MVA peptide pools used in this study are described in Supplementary Table 1.

### IFN-**γ** Elispot assay

Ag-specific T cell responses were assessed using an IFN-γ ELISpot assay (ELISpot plus: Human IFN-γ (ALP), Mabtech) on cryopreserved PBMCs following the manufacturer’s instructions. Thawed PBMCs were stimulated (100,000 cells/well) for 18 h at 37°C, 5% CO_2_ with 2 µg/ml of the 6 PepMix as described previously. Non-stimulated PBMCs were used as a negative control and anti-CD3-(1/2000) stimulated PBMCs were used as a positive control. Spot-forming cells (SFC) were counted using an AID ELISpot reader (AID Autoimmun Diagnostika GmbH) and are expressed as SFC/10^6^ cells after deduction of the background (non-stimulated PBMCs). Seventy-eight samples were tested, and the positivity threshold was defined as three-fold the mean of all non-stimulated PBMCs (52 SFC/10^6^ cells) after deduction of the background.

### Intracellular Staining (ICS) assay

The quality of Ag-specific T-cell responses was evaluated using an intracellular cytokine staining (ICS) assay on cryopreserved PBMCs. PBMCs were stimulated (18h, 37°C, 5% CO_2_) with the six peptide pools as described above in the presence of anti-CD28 and anti-CD49d Abs (1 µg/ml each) and GolgiPlug (10 µg/ml) (BD Biosciences, Le Pont de Claix, France). Cell functionality was assessed by ICS, with Boolean gating. The flow cytometry panel included a viability marker, CD3, CD4, and CD8 antibodies to determine the T-cell lineage, and IFN-γ, TNF, IL-2, IL-4, IL-10, IL-13 antibodies (all from BD Biosciences).

Data were acquired using a LSRFortessa four-laser (488, 640, 561, and 405 nm) flow cytometer (BD Biosciences) and analyzed using FlowJo software, version 9.9.5 (Tree Star). Distributions were plotted with SPICE version 5.22, downloaded from http://exon.niaid.nih.gov/spice ^41^

### Statistical analysis

Figures and statistical analyses were conducted using GraphPad Prism 9. Statistical significance between groups was calculated using the tests indicated in each figure legend. No statistical methods were used to predetermine sample size.

