## Supplemental for "Characterization and comparison of immunity against MPXV for individuals infected with MPXV or vaccinated with modified vaccinia Ankara vaccines"

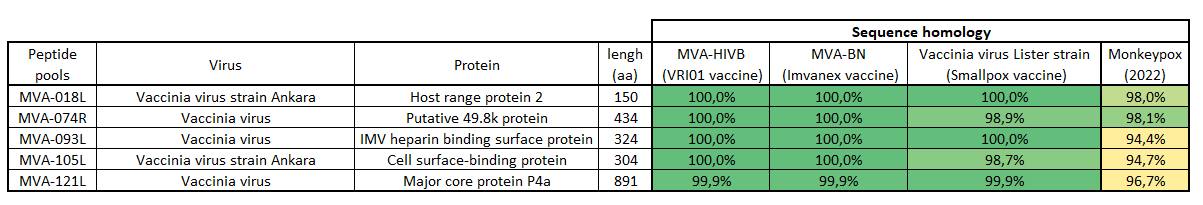

**Supplementary Table 1. Description of the MVA peptide pools used in this study**

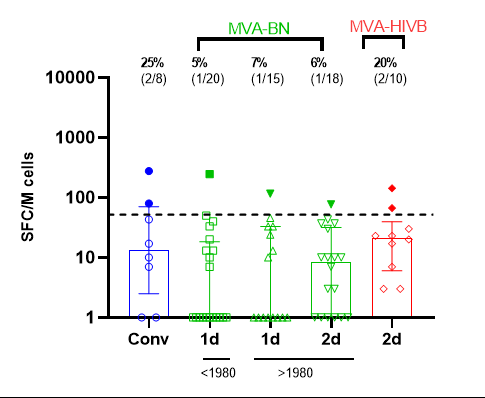

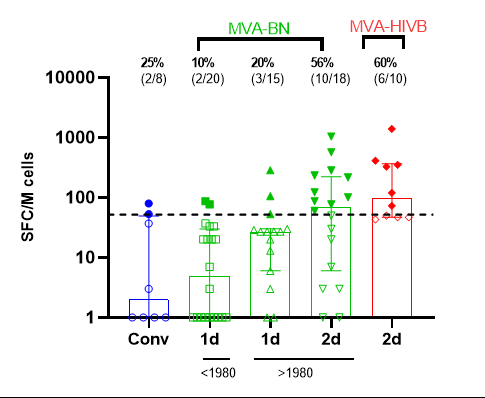

**MVA-074R**

**MVA-018L**

**MVA-0105L**

**MVA-093L**

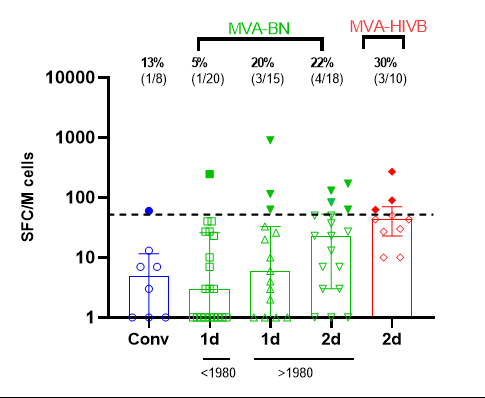

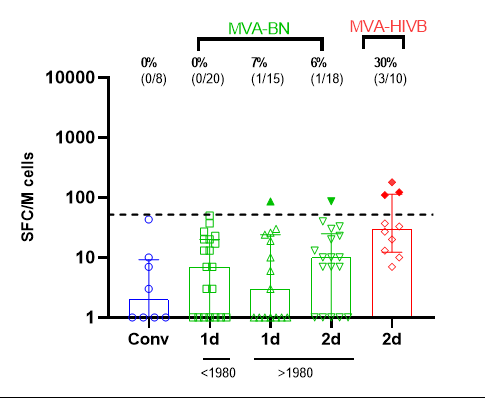

**MVA-121L**

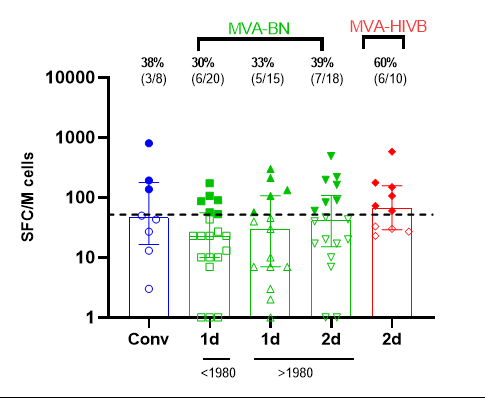

**Supplementary Figure 1. Frequency and magnitude of T-cell responses against individual MVA peptide pools induced by MPXV infection and the MVA-BN and experimental MVA-HIVB vaccines.** Frequency of responders and the magnitude of T-cell responses (SFC/million) of MPXV convalescent patients (Conv) (n=8) and MVA-BN vaccinees (n=20 and n=15 after the first dose (1d) for individuals born <1980 and >1980, respectively, n=18 after the second dose (2d)) and MVA-HIVB vaccinees (n=10) against the MVA-018L, -074R, -093L, -0105L, and -121L peptide pools measured by IFN-γ ELISPOT assay. Background-subtracted results are presented. The dotted line represents the positivity threshold described in the materials and methods section.

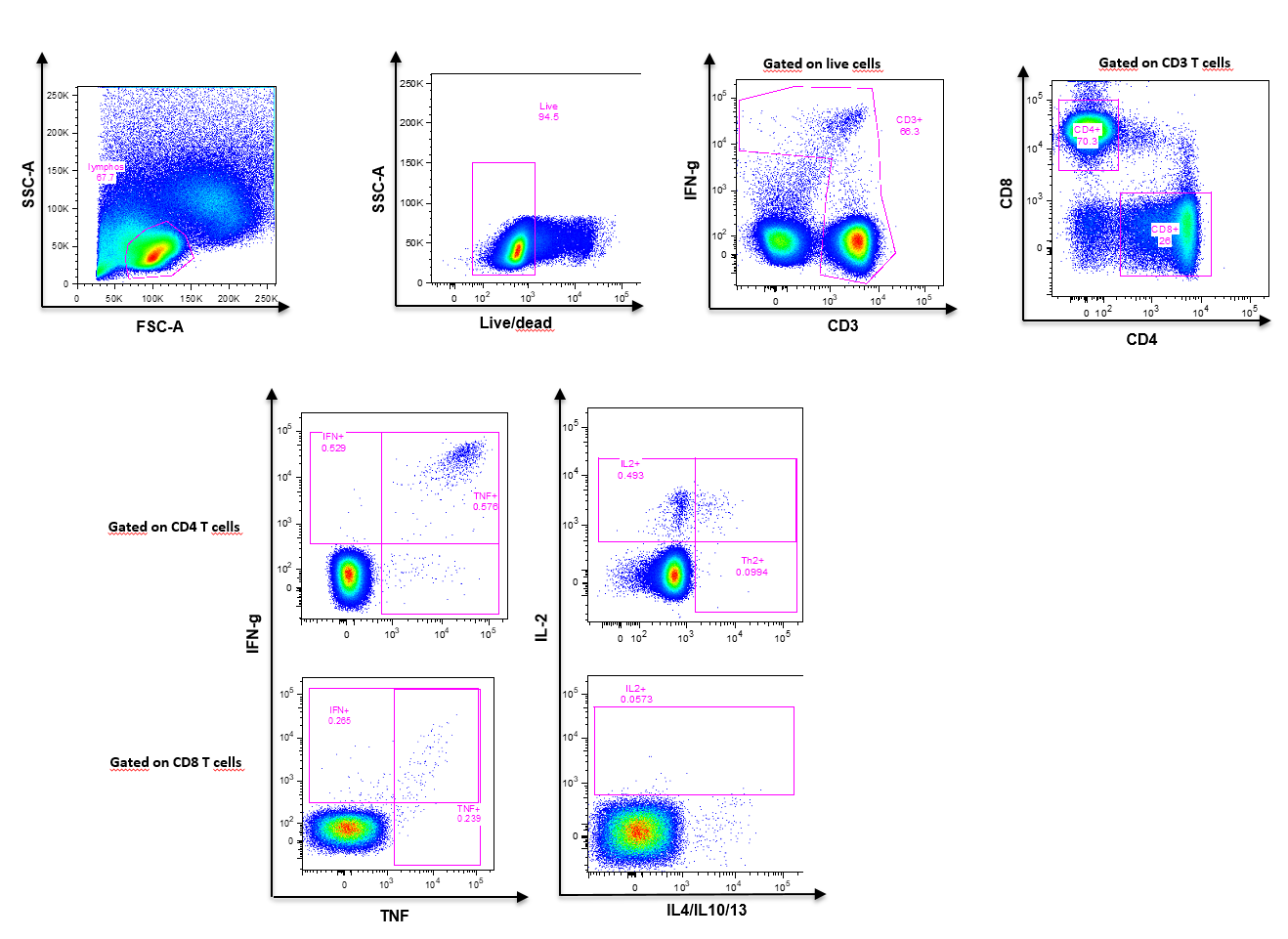

**Supplementary Figure 2:** Gating strategy and representative dot plots of MVA-121L-specific CD4+ and CD8+ T-cell responses after overnight in vitro stimulation of mpox-convalescent PBMCs with MVA-121L peptide pool (2 µg/mL).
